# Disentangling the direct and indirect effects of childhood adiposity on type 1 diabetes and immune-associated diseases: a multivariable Mendelian randomization study

**DOI:** 10.1101/2021.04.19.21255222

**Authors:** Tom G Richardson, Daniel J M Crouch, Grace M Power, Fernanda Morales Berstein, Emma Hazelwood, Si Fang, Yoonsu Cho, Jamie R J Inshaw, Catherine C Robertson, Carlo Sidore, Francesco Cucca, Steven S Rich, John A Todd, George Davey Smith

**Affiliations:** MRC Integrative Epidemiology Unit (IEU), Population Health Sciences, Bristol Medical School, University of Bristol, Oakfield House, Oakfield Grove, Bristol, United Kingdom; Novo Nordisk Research Centre Oxford, Old Road Campus, Oxford, United Kingdom; JDRF/Wellcome Diabetes and Inflammation Laboratory, Wellcome Centre for Human Genetics, Nuffield Department of Medicine, NIHR Biomedical Research Centre, University of Oxford, Oxford, United Kingdom; Center for Public Health Genomics, University of Virginia, Charlottesville, Virginia, USA; Institute for Research in Genetics and Biomedicine (IRGB), Sardinia, Italy

**Keywords:** Childhood adiposity, Type 1 diabetes, Mendelian randomization, chronic immune-associated diseases, inflammation, β-cell fragility

## Abstract

**Background:** The rising prevalence of childhood obesity has been postulated as an explanation for the increasing rate of individuals diagnosed with type 1 diabetes (T1D). However, robust causal evidence supporting this claim has been extremely challenging to uncover, particularly given the typical early onset of T1D.

**Methods:** In this study, we used genetic variation to separate the direct effect of childhood body size on T1D risk from the effects of body size at different stages in the life course using univariable and multivariable Mendelian randomization (MR). Similar MR analyses were conducted on risk of seven other chronic immune-associated diseases.

**Findings:** Childhood body size provided evidence of an effect on T1D (based on a sample of 5,913 cases and 8,282 controls) using a univariable model (OR=2.05 per change in body size category, 95% CI=1.20 to 3.50, P=0.008), which remained after accounting for body size at birth and during adulthood (OR=2.32, 95% CI=1.21 to 4.42, P=0.013). The direct effect of childhood body size was validated using data from a large-scale T1D meta-analysis based on n=15,573 cases and n=158,408 controls (OR=1.94, 95% CI=1.21 to 3.12, P=0.006). We also obtained evidence that childhood adiposity influences risk of asthma (OR=1.31, 95% CI=1.08 to 1.60, P=0.007), eczema (OR=1.25, 95% CI=1.03 to 1.51, P=0.024) and hypothyroidism (OR=1.42, 95% CI=1.12 to 1.80, P=0.004). However, these estimates all attenuated to the null when accounting for adult body size, suggesting that the effect of childhood adiposity on these outcomes is mediated by adiposity in later life.

**Interpretation:** Our findings support a causal role for higher childhood adiposity on higher risk of being diagnosed with T1D. In contrast, the effect of childhood adiposity on the other immune-associated diseases studied was explained by a long-term effect of remaining overweight for many years over the life course.

## Introduction

The incidence of type 1 diabetes (T1D) has doubled in the last 20 years. Possible explanations for this increasing T1D burden include secular changes to gut microbiota linked to the hygiene hypothesis in which increased sanitation^1^, urban living and other factors contribute to increases in not only T1D but in a number of other immune system related diseases, such as multiple sclerosis and asthma^2^. Additional explanations for this increasing burden include the association of virus infection with T1D^3^ and decreasing levels of vitamin D in the population^4^. One hypothesis is that the rising prevalence of childhood obesity in an increasingly obesogenic environment^5-7^, including poor diets with high fat, salt and carbohydrate, may contribute towards early life β-cell fragility and increased susceptibility to T1D^8^. Developing insight into the contribution of childhood adiposity to T1D risk is extremely challenging, however, particularly in terms of separating its effect from early life confounding factors such as birthweight^9^.

In contrast to T1D, there is irrefutable evidence that children who are overweight are more likely to develop type 2 diabetes (T2D) and that weight loss can lead to its sustained remission^10^. We recently used human genetic data to infer that this relationship is likely to be causal rather than due to confounding factors, using sets of genetic variants which robustly associate with childhood and adulthood body size^11^. This was achieved using Mendelian randomization (MR), which can be implemented through an instrumental variable analysis, exploiting the quasi-random assortment of genetic alleles at birth to infer causality between lifestyle exposures and disease outcomes^12-14^.

We showed previously that childhood adiposity increases T2D risk when analysed in a univariable setting (Odds Ratio (OR)=2.32, 95% confidence interval (CI)=1.76 to 3.05, P=3.83×10^−9^) (**Figure 1A**)^11^. However, by simultaneously estimating the genetically predicted effects of childhood adiposity and adulthood adiposity as separate exposures onto T2D risk using a multivariable model, the childhood estimates attenuated to include the null (OR=1.16, 95% CI=0.74 to 1.82, P=0.52). As such, there is considerably weaker evidence that childhood adiposity has a ‘direct effect’ on T2D risk (**Figure 1B**), as compared to it having an ‘indirect effect’ mediated via adult adiposity (**Figure 1C**). These results therefore suggest that the univariable estimates for childhood adiposity can be explained by long term, persistent effects of adiposity due to individuals typically remaining overweight into adulthood.

**Figure 1:**
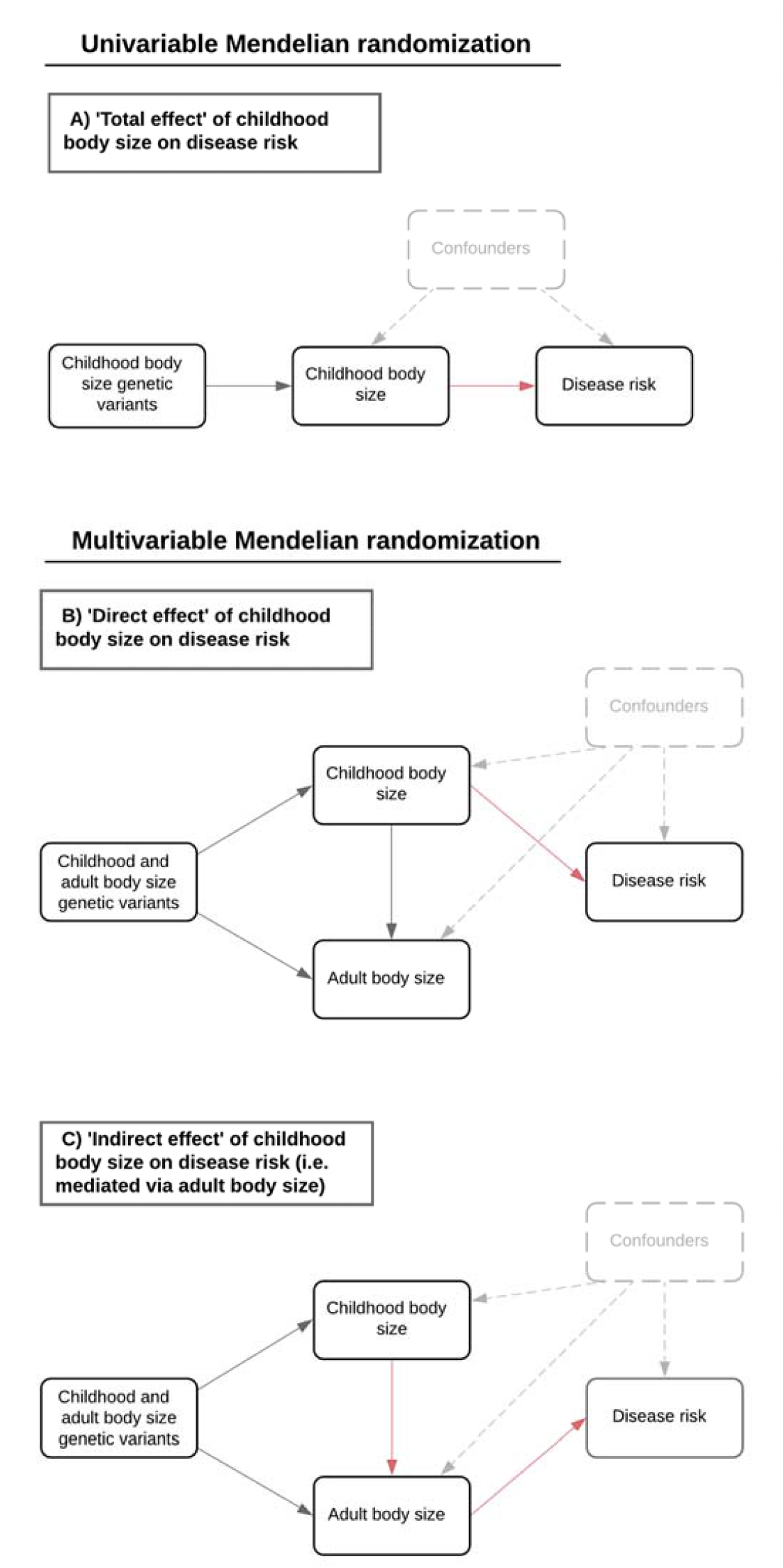
Directed acyclic graphs depicting the effects of childhood adiposity on disease risk.

Although childhood adiposity has been previously implicated in T1D risk using MR^15^, these findings were based on effect estimates derived using a small number of instruments (n=23). Furthermore, multivariable analyses in this study did not model potential confounding factors, such as birthweight, which may be pleiotropically influenced by genetic instruments for the exposure of interest. This is particularly important, as exemplified by the case of high density lipoprotein cholesterol onto coronary heart disease risk, which appears to have a protective effect in a univariable setting (OR=0.80, 95% CI=0.75 to 0.86, P=1.66×10^−10^), but not when assessing its direct effect after taking into account atherogenic lipoprotein lipid traits (OR=0.91, 95% CI=0.74 to 1.12, P=0.36)^16^. Lastly, it has not yet been investigated whether the effect of childhood adiposity on T1D risk represents a more generalizable effect on the immune system which may additionally impact other types of immune-associated or autoinflammatory diseases. If there is a T1D-specific effect, this would suggest early life β-cell fragility stemming from diet-induced metabolic stress is likely to be a causal pathway through which childhood adiposity leads to increased T1D risk.

Consequently, in the present study we had four aims:

1. Investigate evidence of a direct effect of childhood adiposity on T1D risk by conducting univariable and multivariable MR analyses using our previously developed framework (with n=280 genetic instruments).
2. Determine whether these childhood estimates based on age 10 years body size remain robust after accounting for very early life body size as proxied by genetically predicted birthweight.
3. Evaluate the converse relationships using MR i.e. whether T1D genetic liability influences body size in childhood or adulthood.
4. Investigate whether childhood adiposity has direct and indirect effects on seven other types of immune-associated or autoinflammatory diseases.

## Materials and Methods

### Data resources

#### Genetic instruments for childhood and adult body size

Genetic variants associated with childhood and adult body size (based on P<5×10^−8^) were identified from a previously undertaken GWAS in the UK Biobank (UKB) study^17,18^. Analyses have been described in-detail previously^11^. We derived our childhood body size measure using recall questionnaire data asking UKB participants if they were ‘thinner’, ‘plumper’ or ‘about average’ when they were aged 10 years old compared to the average. Adult body size was derived using clinically measured body mass index (BMI) data (mean age 56.5 years), which we categorized into a 3-tier variable using the same proportion as the early life measure for comparative purposes.

GWAS were undertaken on 453,169 individuals who had both measures available with adjustment for age, sex and genotyping chip. Our GWAS of childhood adiposity was additionally adjusted for month of birth. We used a linear mixed model to account for genetic relatedness and geographical structure in UKB as undertaken with the BOLT-LMM software. To support the robustness of these instruments in terms of their ability to separate the effects of childhood and adult body size, we have previously undertaken validation analyses using measured BMI data from three independent populations: the Avon Longitudinal Study of Parents and Children (ALSPAC)^11^, the Young Finns Study^19^ and the Trøndelag Health (HUNT) study^20^. Other validation analyses have also been conducted previously, whereby GWAS results for the childhood measure had a higher genetic correlation with measured childhood obesity from an independent sample (rG=0.85) compared to the adult measure (rG=0.67). Conversely, genome-wide estimates for the adult measure were more strongly correlated with measured BMI in adulthood (rG=0.96) compared to the childhood measure (rG=0.64)^11^. Furthermore, using these instruments previously for multivariable MR provided F-statistics > 10 suggesting that derived results are unlikely to be prone to weak instrument bias^11^.

#### Genetic instruments for childhood height, adult height and birthweight

Here, we repeated the same protocol described above but for childhood and adult height using data from the UKB study, to demonstrate that our body size was likely capturing adiposity rather than being bigger at age 10. Participants were asked “When you were 10 years old, compared to average would you describe yourself as…”, and given the options of ‘shorter’, ‘about average’ or ‘taller’. GWAS were undertaken as above on the childhood measure of height as well as a 3-tiered categorical variable for adult measured height based on the same proportions. GWAS on childhood and adult height were undertaken on 454,023 individuals who had both measures available with adjustment for the same covariates as before. The same analysis pipeline was applied to generate genetic instruments for birthweight on a total of 261,932 UKB individuals. This trait was rank-based inverse normal transformed to ensure normality and adjusted as before for age, sex and genotyping chip.

#### Genetic effects on T1D, T2D and other immune-associated diseases

Genetic estimates for all outcomes analysed in this study were obtained from large-scale GWAS studies and meta-analyses conducted by consortia. We firstly applied our multivariable approach using a large number of childhood and adult body size instruments to T1D data analysed previously in the study by Censin et al. (n=5,913 cases and n=8,828 controls). Results from this analysis were then validated using a recent large-scale meta-analysis of up to 15,573 cases and 158,408 controls^21^. Analyses were then repeated separately in each contributing cohort from this meta-analysis.

We also obtained estimates using results from a GWAS of T2D, updated since our previous study^22^, and seven of the most common immune-associated diseases: asthma, Crohn’s disease, atopic dermatitis and eczema, hypothyroidism, inflammatory bowel disease, rheumatoid arthritis and ulcerative colitis. An overview of these outcome datasets and all others analysed in this study can be found in **Supplementary Table 1**.

#### Instrument identification and data harmonization

We previously constructed a reference panel based using genotype data from 10,000 unrelated UK Biobank participants of European descent to undertake linkage disequilibrium (LD) clumping^23^. This allowed us to identify independent genetic variants for MR analyses based on an LD cutoff of r^2^<0.001^24^, which was necessary to ensure MR estimates were not biased by using correlated instruments. For multivariable MR, we repeated LD clumping but using aggregated sets of genetic variants for all our exposures to ensure they were also independent. Genetic estimates for our exposures were harmonized with disease outcomes using the ‘TwoSampleMR’ R package^25^. In total, there were 280 childhood body size and 515 adult body size instruments, 629 childhood height and 907 adult height instruments and 161 birthweight instruments after harmonization with T1D genetic estimates. The number of instruments for all subsequent analyses varied depending on factors such as coverage, population allele frequencies and the strand alignment of corresponding GWAS results.

### Statistical analysis

#### Univariable Mendelian randomization

We firstly undertook univariable MR analyses to evaluate the total effect of genetically predicted childhood body size on T1D risk. We applied the inverse variance weighted (IVW) method for initial analyses, which takes the SNP-outcome estimates and regresses them on those for the SNP-exposure associations^26^. The weighted median and MR-Egger methods were subsequently applied as sensitivity analyses to evaluate the robustness of IVW estimates to horizontal pleiotropy^27,28^. This is the phenomenon whereby genetic variants influence an exposure and outcome via two separate biological pathways^13^.

Univariable analyses with T1D as an outcome were repeated separately for adult body size and for birthweight. We included adult body size to demonstrate the importance of using genetic scores to separate the effects of adiposity at different stages in the life course when investigating either early or late onset disease outcomes. Additionally, we investigated the opposite direction of effect using the same univariable methods mentioned above to assess whether genetic liability towards T1D risk influences body size in both childhood and adulthood in turn. In this analysis we used a set of genetic instruments for T1D identified from a recent meta-analysis (of up to 15,573 cases and 158,408 controls^21^) and adult BMI was analysed as a continuous trait to derive a per standard deviation effect estimates.

Furthermore, birthweight was only analysed in this study to investigate whether an individual’s body size in very early life (e.g. before age 5 years) may be responsible for the effects identified using our childhood genetic score (**Supplementary Figure 1**). These analyses were not however intended as an exploration of the effects of parental influences on T1D risk^29^, as birthweight variation is known to be influenced by a combination of both fetal and parental genetic and non-genetic factors^30^. We also repeated analyses on T1D using instruments for childhood and adult height to demonstrate that our childhood body size measure was capturing childhood adiposity (i.e. being ‘plumper’ as described in the questionnaire) rather than being taller than other 10-year olds.

#### Multivariable Mendelian randomization

We next sought to estimate the direct and indirect effect of childhood body size on T1D risk using multivariable IVW MR^31,32^. This was firstly undertaken by accounting for adult body size as an additional exposure in our model (i.e. alongside childhood body size), and subsequently including birthweight as a third exposure. We also applied the multivariable MR Egger method to evaluate horizontal pleiotropy for the direct and indirect effects of childhood body size^33^. Furthermore, multivariable analyses were repeated using data from the large-scale T1D meta-analysis^21^, as well as evaluating evidence using data from each contributing cohort to this dataset in turn. Lastly, we repeated our multivariable MR analysis with childhood and adult body size as exposures onto each of the seven different types of immune-associated/autoinflammatory disease in turn. To account for multiple testing in this analysis, we applied the Benjamini-Hochberg false discovery rate (FDR) correction of FDR<5%.

Forest plots in this paper were generated using the R package ‘ggplot2’^34^. All analyses were undertaken using R (version 3.5.1).

## Results

### Estimating the total effect of childhood adiposity on type 1 diabetes risk

Univariable MR analyses using the IVW method provided evidence that both childhood adiposity (Odds Ratio (OR)=2.05 per change in body size category, 95% confidence interval (CI)=1.20 to 3.50, P=0.008) and adult adiposity (OR=1.60, 95% CI=1.05 to 2.45, P=0.03) increase risk of T1D. The total effect of childhood body size was additionally supported by the MR-Egger method (OR=5.06, 95% CI=1.52 to 16.81, P=0.009), suggesting that this result is robust to horizontal pleiotropy. In contrast, we obtained no convincing support that adult adiposity influences T1D based on the MR-Egger method (OR=2.55, 95% CI=0.72 to 9.00, P=0.145) (**Supplementary Table 2**).

Repeating our univariable IVW analysis using childhood and adult height instruments provided no support of effects on T1D (childhood height: OR=1.16, 95% CI=0.94 to 1.44, P=0.174, adult height: OR=1.08, 95% CI=0.85 to 1.36, P=0.532) (**Supplementary Table 3**). These findings provide evidence that our estimates for childhood body size on T1D are capturing an adiposity driven effect as opposed to a general body size effect. Furthermore, evidence of a total effect between childhood body size on T1D risk was identified in the largest available T1D meta-analysis to date (IVW: OR=1.84, 95% CI=1.19 to 2.83, P=0.006, MR-Egger: OR=3.28, 95%=1.24 to 8.67, P=0.017) (**Supplementary Table 4**).

We also identified limited evidence of a converse direction of effect between T1D genetic liability and childhood adiposity (Beta=0.001, 95% CI=-0.002 to 0.004, P=0.620), meaning that the effect of childhood adiposity on T1D is unlikely to be explained by reverse causality. There was evidence, however, to suggest that T1D genetic liability may have an effect on lower BMI in adulthood using the pleiotropy robust MR methods (weighted median: Beta=-0.007 per standard deviation change in BMI, 95% CI=-0.013 to −0.002, P=0.009, MR-Egger: Beta=-0.017, 95% CI=-0.029 to −0.004, P=0.009), although not using the IVW approach (Beta=9.66×10^−5^, 95% CI=-0.006 to 0.006, P=0.975) (**Supplementary Table 5**).

### Evaluating the direct and indirect effects of childhood adiposity on type 1 diabetes risk

Multivariable MR provided evidence that childhood adiposity has a direct effect on T1D risk (OR=2.27, 95% CI=1.24 to 4.17, P=0.008), whereas adult estimates identified in this analysis included the null (OR=0.92, 95% CI=0.54 to 1.57, P=0.760) (**Supplementary Table 6**). Using the multivariable MR-Egger method supported evidence of a direct effect for childhood adiposity on T1D risk (OR=2.20, 95% CI=1.20 to 4.05, P=0.011) (**Supplementary Table 7**).

Repeating our multivariable MR analyses on T1D risk with the addition of genetically predicted birthweight in the model found that the childhood adiposity estimates were maintained (OR=2.32, 95% CI=1.21 to 4.42, P=0.013) (**Supplementary Table 8**). Additionally, higher genetically predicted birthweight provided evidence of a protective direct effect on T1D risk (OR=0.58, 95% CI=0.41 to 0.82, P=0.002) independent of childhood and adult body size. These results suggest that body size at birth is unlikely to be responsible for the effect of childhood adiposity on T1D in our model. Furthermore, univariable estimates for birthweight on T1D risk were not robust to horizontal pleiotropy based on estimates from the MR-Egger method (OR=0.44, 95% CI=0.16 to 1.24, P=0.124) (**Supplementary Table 9**). Multivariable MR estimates for adult body size on T1D, accounting for genetically predicted birthweight, did not support a role for obesity later in life influencing T1D (OR=0.77, 95% CI=0.43 to 1.39, P=0.390).

Evidence of a direct effect between childhood adiposity and T1D risk was validated using data from the large meta-analysis of T1D GWAS (OR=1.94, 95% CI=1.21 to 3.12, P=0.006) (**Supplementary Table 10**). Direct effect estimates derived from each contributing dataset to the T1D meta-analysis were typically consistent with the exception of the cohort from Sardinia (**Supplementary Figures 2-4 & Supplementary Table 11**). We also repeated our multivariable MR analysis of childhood and adult body size with T2D as an outcome to generate revised estimates compared to our previous work. In contrast to our results for T1D, these estimates suggest that childhood adiposity has an indirect on T2D as our univariable childhood estimates (OR=2.18, 95% CI=1.80 to 2.63, P=8.91×10^−16^) were reduced and included the null when accounting for adult body size (OR=0.90, 95% CI=0.69 to 1.19, P=0.465) (**Supplementary Table 12**). Forest plots illustrating the univariable and multivariable estimates for all analyses in this section can be found in **Figure 2**.

**Figure 2:**
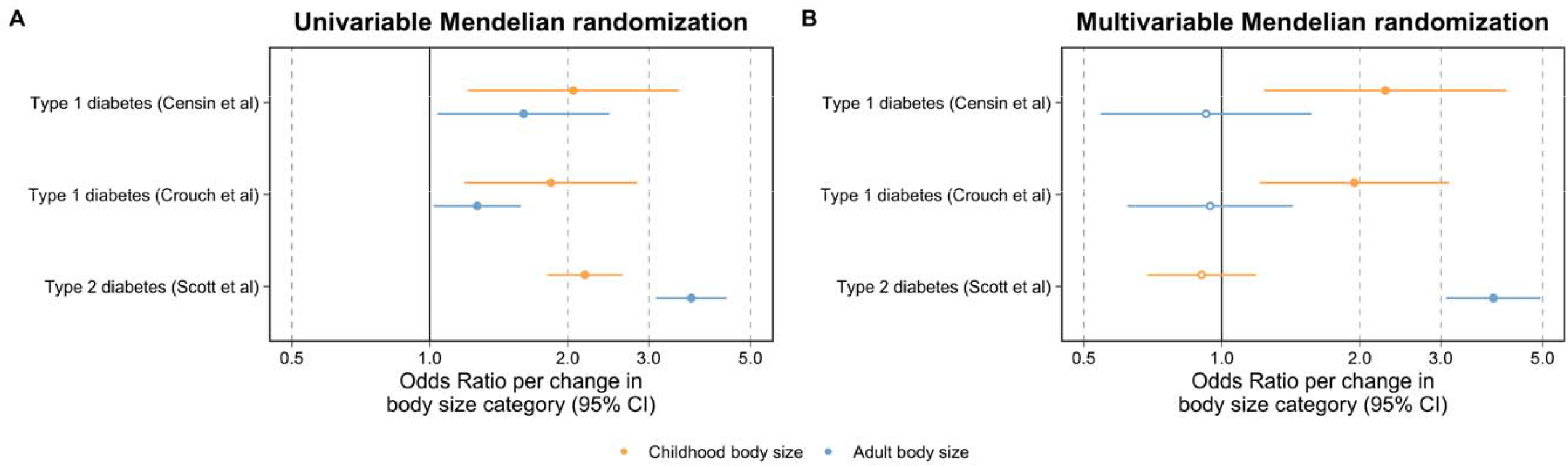
Forest plots illustrating the univariable and multivariable Mendelian randomization estimates of childhood adiposity on type 1 and type 2 diabetes risk. A) The univariable Mendelian randomization (MR) estimates between childhood (yellow) and adult (blue) body size on risk of type 1 (using estimates from both discovery and replication analysis) and type 2 diabetes and B) their corresponding multivariable MR estimates. Odds ratios are per change in body size category. 95% CI = 95% confidence interval.

### Investigating whether childhood adiposity directly influences other types of immune disease

We applied univariable and multivariable MR analyses to each of the seven immune-associated diseases in turn. Using univariable MR, 10 of the 16 analyses undertaken provided evidence that adiposity in either childhood or adulthood influenced chronic immune disease risk based on FDR<5% (**Supplementary Table 13**). For childhood adiposity, this included evidence of increased asthma risk (OR=1.31, 95% CI=1.08 to 1.60, P=0.007), dermatitis and eczema (OR=1.25, 95% CI=1.03 to 1.51, P=0.024) and hypothyroidism (OR=1.42, 95% CI=1.12 to 1.80, P=0.004). Adult adiposity provided evidence of influencing risk on outcomes including Crohn’s disease (OR=1.37, 95% CI=1.10 to 1.70, P=0.005) and rheumatoid arthritis (OR=1.42, 95% CI=1.05 to 1.93, P=0.022).

Using multivariable MR, the direct effect estimates for childhood adiposity, on all immune-associated and autoinflammatory disease outcomes which provided evidence of an effect in a univariable setting, included the null when accounting for the effect of adult adiposity (**Supplementary Table 14**). We did however identify evidence that childhood adiposity indirectly influences disease risk via adult body size; for asthma risk (OR=1.34, 95% CI=1.15 to 1.57, P=1.93×10^−4^), dermatitis and eczema (OR=1.33, 95% CI=1.12 to 1.57, P=9.04×10^−4^) and hypothyroidism (OR=1.91, 95% CI=1.56 to 2.34, P=4.47×10^−10^). All univariable and multivariable MR estimates derived in these analyses have been illustrated using forest plots in **Figure 3**.

**Figure 3:**
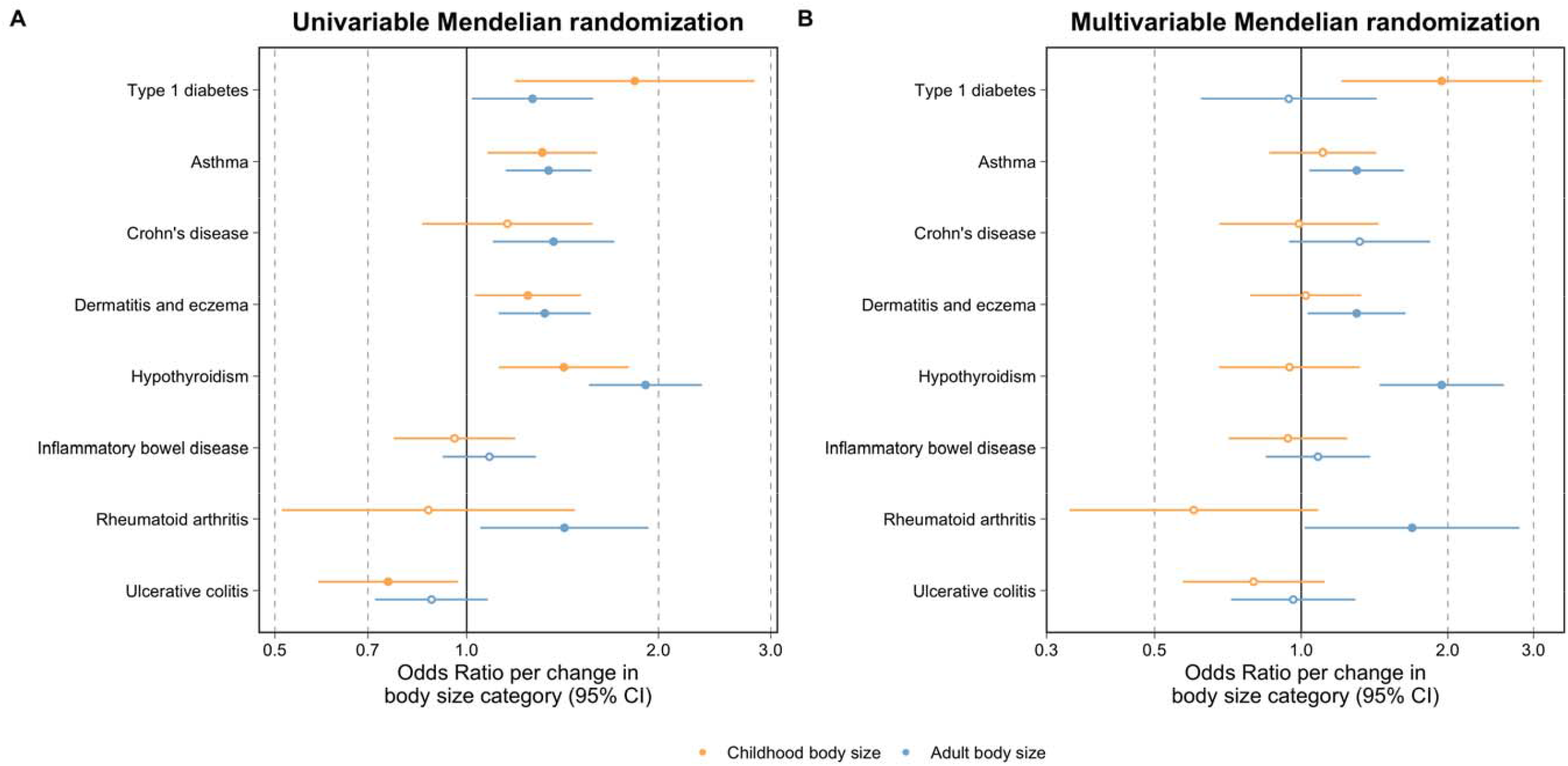
Forest plots comparing the univariable and multivariable Mendelian randomization estimates of childhood adiposity on type 1 diabetes risk and seven chronic immune-associated disease outcomes. A) The univariable Mendelian randomization (MR) estimates between childhood (yellow) and adult (blue) body size on risk of chronic immune disease outcomes and B) their corresponding multivariable MR estimates. The type 1 diabetes estimates were based on the analysis using data from Crouch et al (2021). Odds ratios are per change in body size category. 95% CI = 95% confidence interval.

## Discussion

We present evidence that adiposity in childhood increases the risk of T1D independently of body size at birth and adulthood. These findings support previous results from observational studies suggesting that the increasing prevalence of childhood obesity is a causal factor in the rising numbers of T1D diagnoses. Systematically applying our MR framework to seven other immune-associated diseases suggested, initially, that childhood adiposity also increases risk of asthma, eczema and hypothyroidism. However, these effect estimates attenuated once accounting for adulthood body size, suggesting that they can be explained due to the sustained impact of adiposity among children who are overweight and thus tend to remain so into adulthood.

The effect of genetically predicted childhood adiposity on T1D risk could have various explanations. For instance, this evidence may support findings from the literature suggesting that excess fat tissue has a deleterious influence on the body’s immune system, potentially with secreted adipokines playing a mediatory role^35^. As outlined by the ‘accelerator hypothesis’^36^, increased stress on insulin demands in children with obesity may contribute to earlier β-cell failure and subsequently an earlier diagnoses of T1D^37^. Evidence from a mouse model of non-immune diabetes induced by a high fat diet indicated that diabetes can result from β-cell fragility^38^, including genetically lower expression of the transcription factor gene, *GLIS3*, which is known to be associated with susceptibility to both T1D and T2D^39^. High fat and carbohydrate diets with low fibre in early life, resulting in childhood obesity, could compromise the metabolic and immune functions of the gut microbiome, where microbiota dysbiosis has been associated with both T2D^40^ and T1D^41^. Regardless of the underlying mechanisms, our findings suggest that a critical window exists in childhood to mitigate the influence of adiposity on the escalating numbers of T1D diagnoses.

As expected given the average age-at-diagnosis of T1D, the effect of childhood body size remained robust after accounting for adult body size using a much larger number of genetic instruments than previously used (n=280 in this study versus n=13 previously^15^). Furthermore, our childhood estimates remained strong even after accounting for birthweight. However, estimates derived using the MR-Egger method only supported the childhood body size effect (OR=5.06, 95% CI=1.52 to 16.81, P=0.009), whereas confidence intervals for both birthweight and adult body size overlapped with the null, suggesting that they may be prone to horizontal pleiotropy.

In particular, the multivariable MR estimates for adult body size illustrate the importance of using our approach to separate the effects of adiposity at separate stages in the life course. This is because the univariable MR estimates for adult body size on their own could conceivably be interpreted as evidence that it influences T1D risk, which is unlikely given the age of onset for this disease. However, taken together with the MR-Egger estimates, our multivariable analysis suggested that this is indeed the case. Whilst we did not find evidence that genetic liability towards T1D may influence childhood adiposity, our results suggest that it may have an effect on lower body size in adulthood based on the MR-Egger and weighted median methods. Medical practitioners promote healthy living among T1D patients in order to keep HbA1c levels low, which is one possible explanation for this result. Further work is required to investigate this finding using age at diagnosis data once it becomes available in large sample sizes, particularly given the challenges of T1D diagnosis in adulthood^42^.

We incorporated birthweight as an additional exposure in our multivariable model to assess whether it may help explain effect of childhood body size on T1D. As our estimates remained robust, these findings do not seem to suggest that variation in birthweight is responsible for the effect of genetically predicted childhood body size on T1D risk identified in our analysis. However, a more appropriate evaluation of the role of birthweight on T1D risk requires in-depth evaluation using both maternal and fetal genetic effects, as undertaken previously, once sample sizes of both maternal and offspring T1D cases are sufficient^30,43^.

Our MR analysis on other types of immune-associated disease suggested that the childhood adiposity effect on T1D is not generalizable to other types of chronic immune disease.

Amongst this finding was evidence of a total effect of childhood adiposity on later life asthma risk which corroborated recent MR results suggesting that increased asthma risk is likely explained by individuals remaining overweight into adulthood^44^. However, our univariable results provide stronger evidence than previously reported that the effect of adiposity on asthma risk begins in childhood, which may potentially be explained by the influence of excess abdominal fat driving systemic inflammation^45^. In particular, our findings suggest that adiposity begins to exert its effect on risk of eczema and hypothyroidism in childhood, which has previously been reported in the literature by non-genetic studies^46,47^. The attenuation of the childhood estimates on these outcomes in our multivariable model suggests that adiposity influences their risk due to a sustained effect of remaining overweight for many years across the life course (similar to our findings for T2D^48^). Further research is therefore necessary to verify whether lifestyle changes enforced post-childhood can alleviate the detrimental effect of childhood adiposity on these outcomes as with T2D^10^. Furthermore, if this is the case then extensive research into the critical windows where this effect begins to become immutable will be extremely important to identify for disease prevention purposes.

There are various strengths and limitations of our study which should be taken into account when interpreting its findings. Firstly, the use of genetic variation in a two-sample MR framework allowed us to analyse a large number of genetic instruments from the UK Biobank sample for body size (n=454,023) with a meta-analysed sample of T1D cases (up to n=15,573), almost twice the number of cases used in a previous study^15^. As such our results are less prone to bias attributed to reverse causation and confounding factors compared to more traditional epidemiology approaches. Furthermore, this study design allowed us to investigate the direct and indirect effects of childhood adiposity of seven other chronic immune diseases which would be extremely challenging to undertake without the use of human genetics. Conversely, one of the major limitations of this work is that our 280 genetic instruments for childhood body size were derived using recall data which may be more prone to bias due to factors such as measurement error. That said, previously conducted simulations and extensive validation studies in three separate populations^11,20,49^ support the use of these instruments to separate the effect of childhood adiposity using these instruments from that of adulthood adiposity.

In conclusion, our findings emphasise the importance of implementing preventative policies to lower the prevalence of childhood obesity and its subsequent influence on the rising numbers of T1D cases. This will help ease healthcare burdens and also potentially improve the quality of life for individuals living with this lifelong disease.

## Data Availability

The availability of all data analysed in this study has been referenced throughout the manuscript and supplementary materials.

## Acknowledgements

We thank the authors and GWAS consortia who made their summary statistics available for the benefit of this study. This work was supported by the Integrative Epidemiology Unit which receives funding from the UK Medical Research Council and the University of Bristol (MC_UU_00011/1). GDS conducts research at the NIHR Biomedical Research Centre at the University Hospitals Bristol NHS Foundation Trust and the University of Bristol. The views expressed in this publication are those of the author(s) and not necessarily those of the NHS, the National Institute for Health Research or the Department of Health. TGR was a UKRI Innovation Research Fellow (MR/S003886/1) whilst undertaking this project. GMP is supported by grant MR/N0137941/1 for the GW4 Biomed Doctoral Training Programme, awarded to the Universities of Bath, Bristol, Cardiff and Exeter from the Medical Research Council (MRC)/UKRI. SF and FMB are supported by Wellcome Trust PhD studentships in Molecular, Genetic and Lifecourse Epidemiology [108902/Z/15/Z & 218495/Z/19/Z respectively].

The work of DJMC, JRJI and JAT was supported by the JDRF [9-2011-253], [5-SRA-2015-130-A-N], [4-SRA-2017-473-A-N]; the Wellcome [091157/Z/10/Z], [107212/Z/15/Z]; [203141/Z/16/Z]. No funding bodies had any role in study design, data collection and analysis, decision to publish, or preparation of the manuscript. Computation used the Oxford Biomedical Research Computing (BMRC) facility, a joint development between the Wellcome Centre for Human Genetics and the Big Data Institute supported by Health Data Research UK and the NIHR Oxford Biomedical Research Centre. Financial support was provided by the Wellcome Trust Core Award Grant Number 203141/Z/16/Z. The views expressed are those of the author(s) and not necessarily those of the NHS, the NIHR or the Department of Health

## Competing interests

JAT is a member of a Human Genetics Advisory Board of GSK. TGR is employed part time by Novo Nordisk outside of this work. All other authors declare no conflict of interest.

## Figure titles and legends

**Figure 1: Directed acyclic graphs depicting the effects of childhood adiposity on disease risk**

Schematic representation of the analysis undertaken in this study using Mendelian randomization (MR) A) Using univariable MR to estimate the total effect of genetically predicted childhood adiposity on type 1 diabetes (T1D) risk without accounting for adulthood adiposity B) Applying multivariable MR to estimate the direct effect of genetically predicted childhood adiposity on T1D risk whilst accounting for the effect of adult adiposity and C) using the same approach to estimate the indirect effect of childhood adiposity of T1D (via adult adiposity). The highlight red lines indicate the causal pathway being evaluated in MR to estimate the A) total, B) direct and C) indirect effects of childhood body size on T1D risk.

**Figure 2: Forest plots illustrating the total and direct effects of childhood adiposity on type 1 and type 2 diabetes risk**

A) The univariable Mendelian randomization (MR) estimates between childhood (yellow) and adult (blue) body size on risk of type 1 (using estimates from both discovery and replication analysis) and type 2 diabetes and B) their corresponding multivariable MR estimates. Odds ratios are per change in body size category. 95% CI = 95% confidence interval.

**Figure 3: Forest plots comparing the univariable and multivariable Mendelian randomization estimates of childhood adiposity on type 1 diabetes risk and seven chronic immune disease outcomes**

A) The univariable Mendelian randomization (MR) estimates between childhood (yellow) and adult (blue) body size on risk of chronic immune disease outcomes and B) their corresponding multivariable MR estimates. Odds ratios are per change in body size category. 95% CI = 95% confidence interval.

## Notes

### Author Declarations

All analyses conducted in this study are based on data from summary-level datasets (whose ethical approval can be found in the corresponding studies referenced) or the UK Biobank study (Research Ethics Committee approval number: 11/NW/0382, app #15825).

