## Supplementary Figures for "Disentangling the direct and indirect effects of childhood adiposity on type 1 diabetes and immune-associated diseases: a multivariable Mendelian randomization study"

**Supplementary Figure 1:**

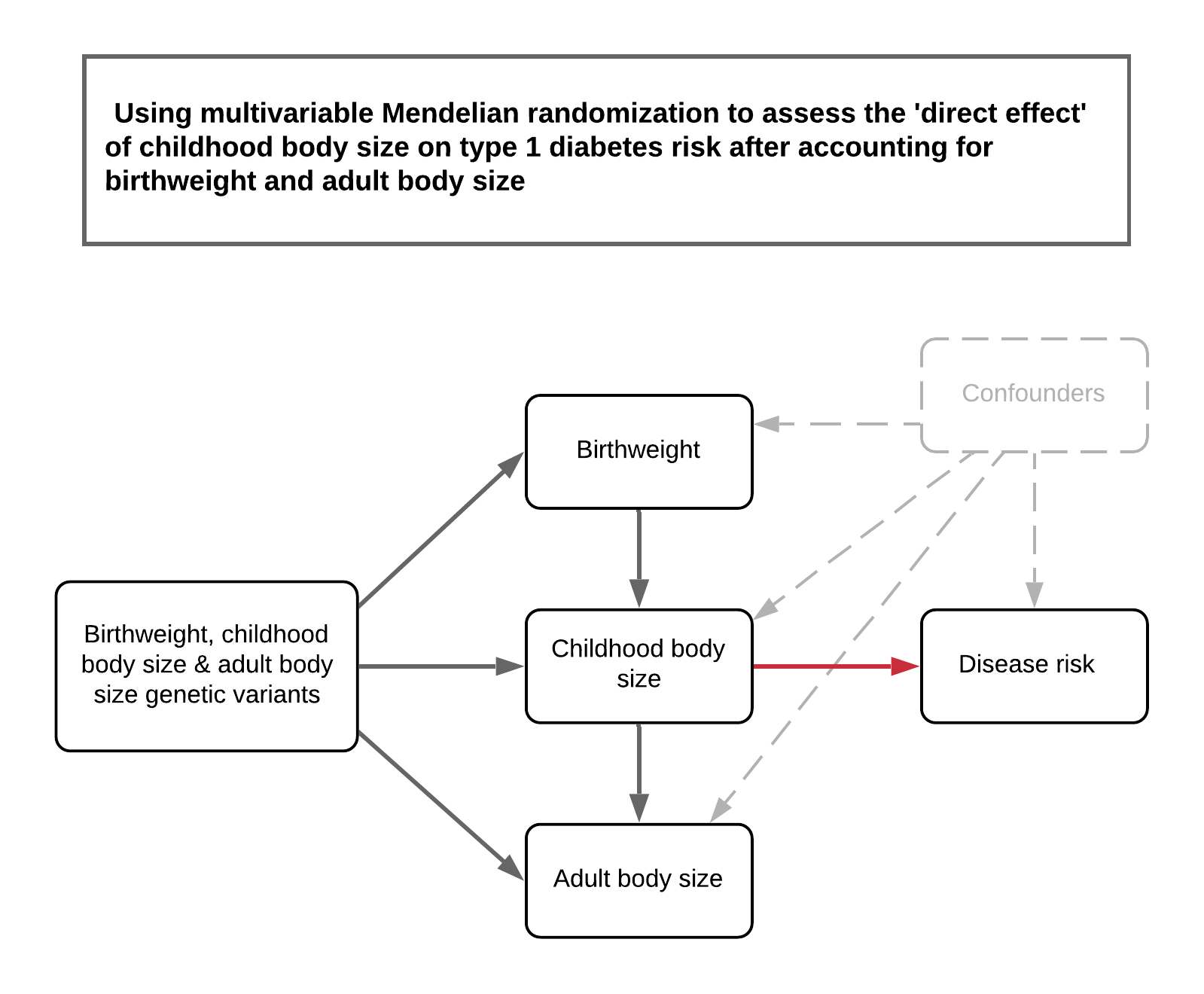

*A schematic representation of the analysis undertaken in this study to assess the direct effect of childhood body after accounting for both birthweight and adult body size using multivariable Mendelian randomization. We emphasise that this analysis was undertaken to investigate whether birthweight was potentially responsible for the direct effect of childhood adiposity, as opposed to rigourously exploring whether parental effects influence type 1 diabetes risk.*

**Supplementary Figure 2:**

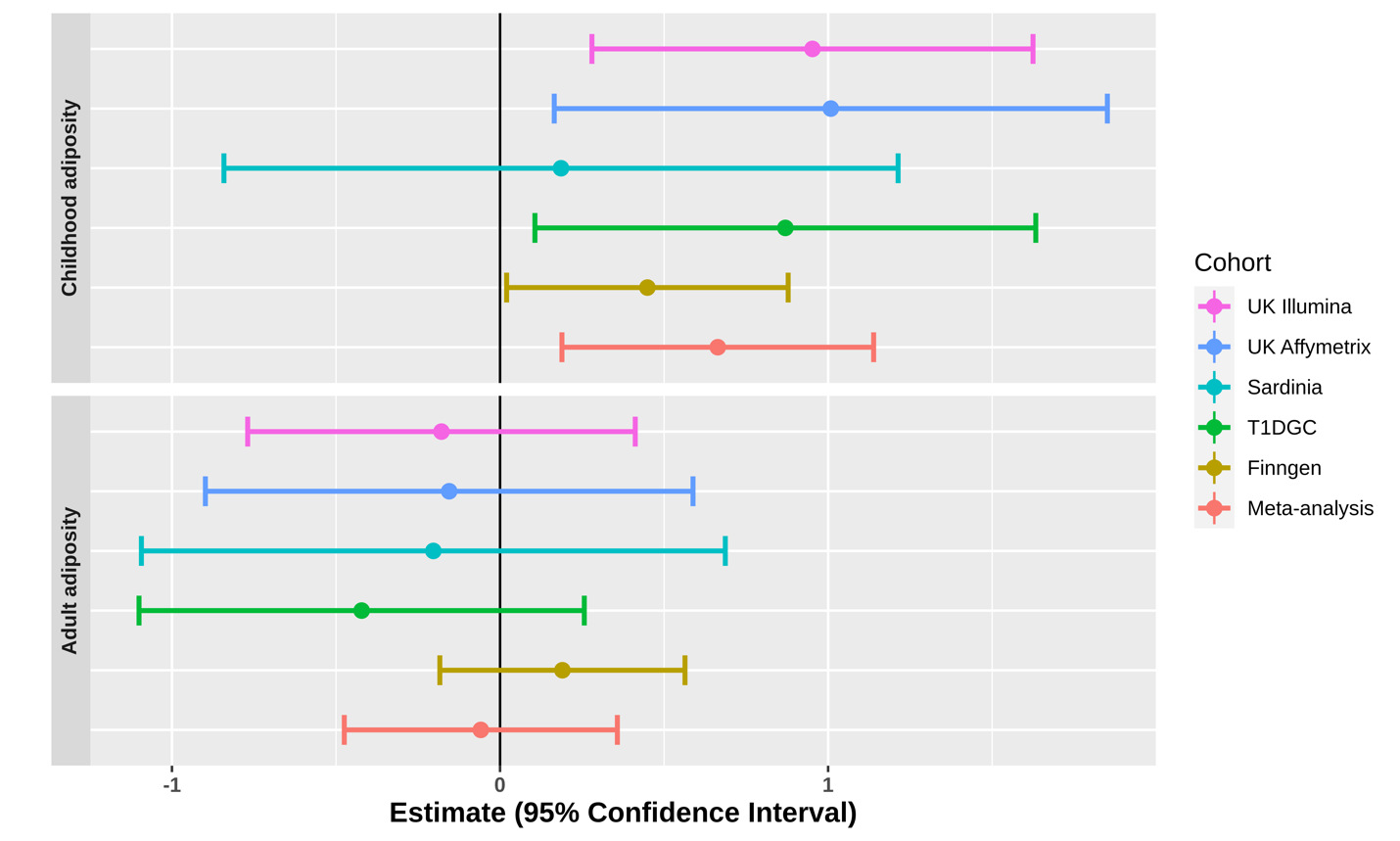

*Multivariable Mendelian randomization analyses of childhood and adult adiposity on type 1 diabetes risk undertaken on each contributing study to the large-scale meta-analysis used in this work. Estimates are based on the inverse variance weighted (IVW) method.*

**Supplementary Figure 3:**

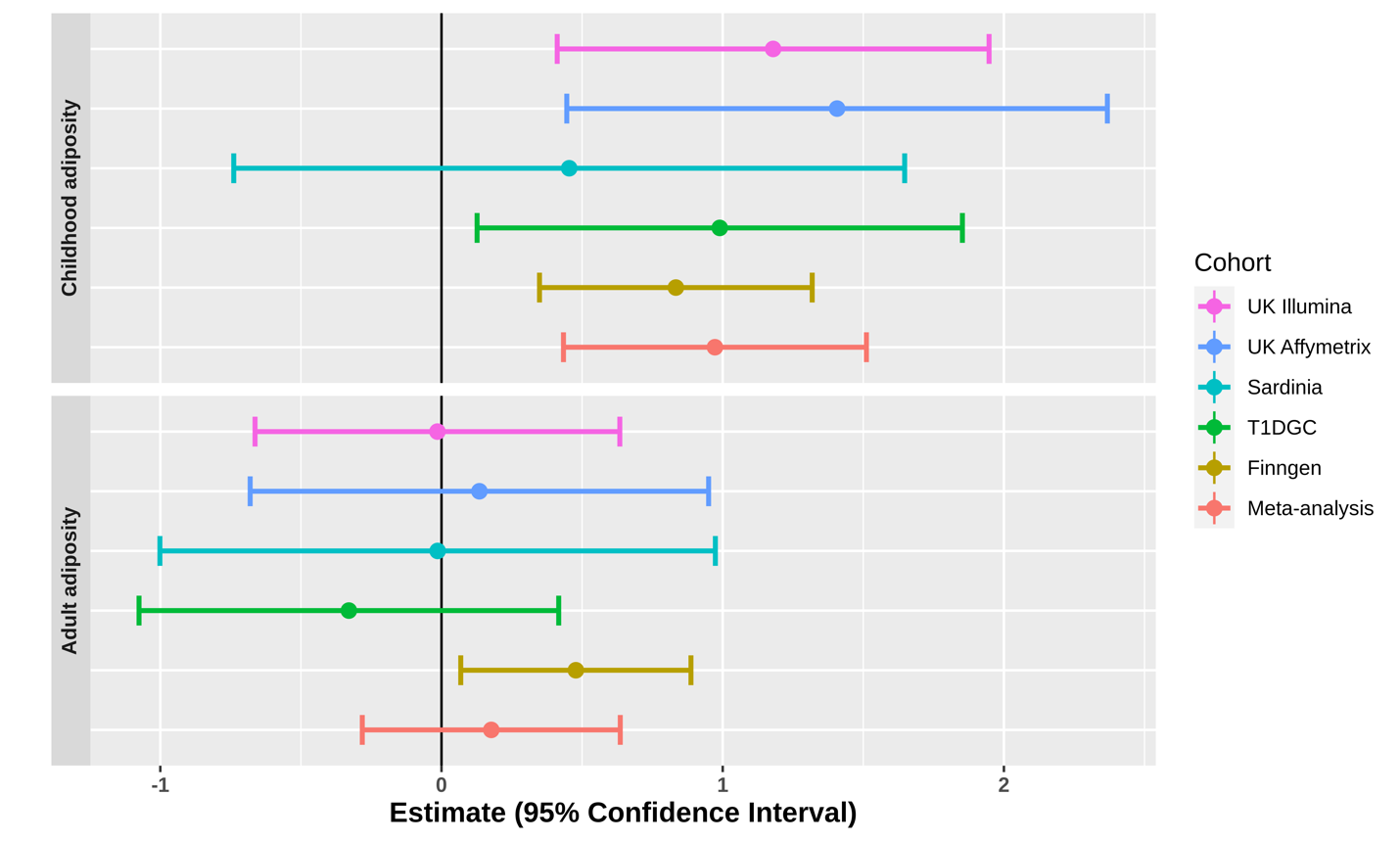

*Multivariable Mendelian randomization analyses of childhood and adult adiposity on type 1 diabetes risk undertaken on each contributing study to the large-scale meta-analysis used in this work. Estimates are based on the MR-Egger method.*

**Supplementary Figure 4:**

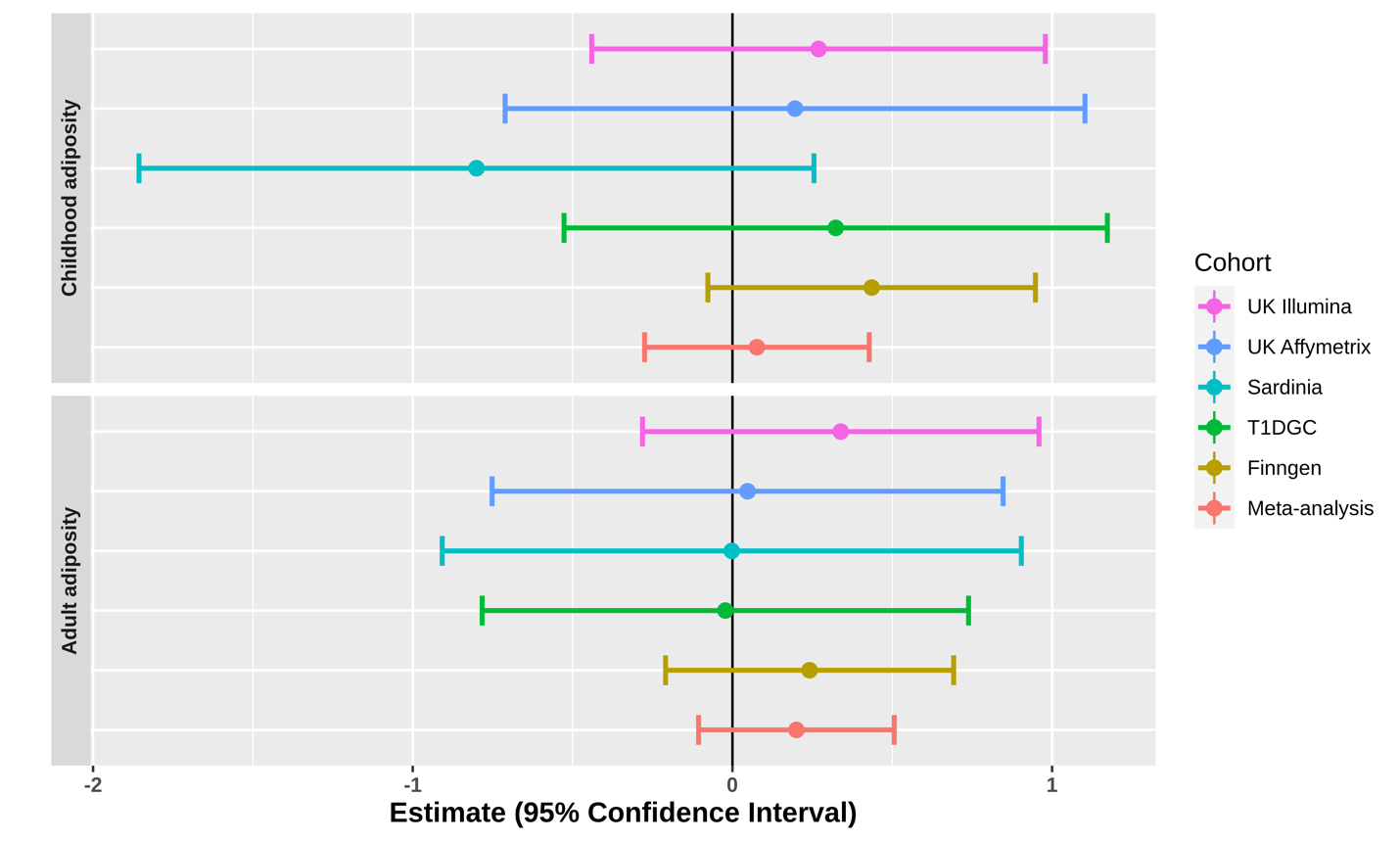

*Multivariable Mendelian randomization analyses of childhood and adult adiposity on type 1 diabetes risk undertaken on each contributing study to the large-scale meta-analysis used in this work. Estimates are based on the weighted median method.*
